# Geospatial evidence of introduction of highly pathogenic avian influenza into dairy farms in the United States

**DOI:** 10.1101/2024.05.02.24306785

**Authors:** Mehak Jindal, Haley Stone, Samsung Lim, Rebecca Dawson, Ashley Quigley, Matthew Scotch, C. Raina MacIntyre

## Abstract

The emergence of Highly Pathogenic Avian Influenza (HPAI) A/H5N1 clade 2.3.4.4b has led to unprecedented intercontinental spread spread across avian and mammalian species. First confirmed outbreaks in dairy farms happened in the United States in March 2024. This study evaluated the modes of introduction and spread across 42 confirmed dairy farm outbreaks spanning nine states between 10 March and 5 May 2024, through geospatial mapping, case-control ecological exposure analysis, and space-time cluster testing. Non-waterfowl abundance was significantly higher in outbreak counties than in high-cattle-density non-outbreak counties (p<0.00001, r=−0.542), identifying it as a distinguishing ecological feature of early dairy outbreak locations. Knox space-time cluster analysis confirmed non-random transmission dynamics across 14 of 30 threshold combinations, consistent with a single introduction in Texas followed by regional and trade-mediated amplification, with livestock trade, poultry litter, and contaminated milking equipment as likely contributing factors. Proactive surveillance targeting counties with high non-waterfowl abundance and cattle density is essential to reduce the risk of further spillover events.

## 1. Introduction

Highly Pathogenic Avian Influenza (HPAI) virus A/H5N1 clade 2.3.4.4b has undergone unprecedented intercontinental spread, affecting a wider range of wild birds and mammalian species than previously recorded (Jindal et al., 2025). Traditionally, avian influenza virus (AIV) spread along migratory routes of wild Anseriformes (waterfowl) before infecting farmed poultry. With the emergence of clade 2.3.4.4b, however, 363 avian species, many non-waterfowl, and 72 mammalian species (aquatic and terrestrial) have now been affected, opening new avenues for transmission (Food and Agriculture Organization of the United Nations, 2024).

Since its first confirmed detection in dairy cattle the United States (US) on 25 March 2024, HPAI A/H5N1 clade 2.3.4.4b has shifted from an unusual cross-species spillover event to an ongoing livestock, wildlife, food-safety, and public-health surveillance problem. The US Department of Agriculture – Animal and Plant Health Inspection Service (USDA APHIS) continues to maintain state-level livestock case surveillance, while Centres for Disease Control and Prevention (CDC) describes A/H5 bird flu as widespread in wild birds, causing outbreaks in poultry and dairy cows, with sporadic human infections among exposed dairy and poultry workers (USDA, 2026). Genomic and epidemiological studies indicate a single spillover into cattle followed by cattle-associated spread, including interstate dissemination linked to dairy production systems (Caserta et al., 2024; USDA, 2025b; Nguyen et al., 2025), with mutations enhancing polymerase activity and the internal gene constellation of genotype B3.13 supporting efficient replication and modulation of bovine antiviral responses (Caserta et al., 2024; Turnbull et al., 2025). However, while these studies have clarified evolutionary relationships and likely introduction events, the geographic conditions that made specific dairy regions vulnerable remain unclear, particularly in distinguishing ecological seeding risk from subsequent spatial amplification via cattle movement, shared farm practices, poultry interfaces, and local transmission.

Control strategies depend on understanding how ecological exposure and farm-associated spread interact across space: if wildlife exposure is the major contributor, surveillance should prioritise interfaces where cattle density overlaps with infected bird abundance; if post-introduction amplification dominates, movement networks, farm connectivity, and poultry-dairy interfaces become the key intervention points (USDA, 2025a). A spatially explicit analysis can therefore provide more actionable evidence than a descriptive outbreak chronology alone.

In this study, we use early confirmed dairy outbreaks as a natural window into the initial spatial structure of A/H5N1 emergence in the United States. We combine county-level dairy outbreak data, cattle density, wild-bird abundance, terrestrial mammal habitat suitability, poultry outbreak proximity, and Knox space-time clustering to test whether early outbreak counties had distinctive ecological exposure profiles and whether outbreak timing showed evidence of non-random space-time structure. Rather than assigning a single transmission route, this approach evaluates whether the observed outbreak geography is more consistent with spatially structured ecological and agricultural risk than with a purely random distribution of cases.

## 2. Methodology

This section outlines the data sources, geospatial methods, and statistical analyses used to investigate the routes of introduction and transmission dynamics of A/H5N1 clade 2.3.4.4b in the US dairy farms between March and 5 May 2024.

### 2.1. Data

#### 2.1.1. Outbreak data

A comprehensive list of all confirmed early H5N1 outbreaks in dairy farms, poultry, domestic animals, wild birds and mammals in the United States from 2021 to 5 May 2024 was compiled from the World Animal Health Information System (WAHIS) (WOAH, 2025). 42 confirmed dairy farm outbreaks were extracted, spanning 10 March to 5 May 2024, filtered to include county-specific location, confirmed start date, and species. A separate list of poultry and domestic animal outbreaks in 2024 was created to assess temporal and spatial proximity to dairy farm cases.

#### 2.1.2. Wild bird abundance data

Estimated weekly relative abundance data were obtained from eBird Status and Trends (Fink et al., 2023) for each wild bird species confirmed to have tested positive for H5N1 in the US, representing modelled mean relative abundance per 14 km^2^ grid cell across the contiguous US for each week of the year. Species were classified as waterfowl (n=36 species) or non-waterfowl (n=103 species) by taxonomic order. Seven species were excluded due to insufficient abundance model coverage: Black-billed Magpie, Black-legged Kittiwake, Brandt’s Cormorant, Gannet, Glaucous Gull, Northern Fulmar, and Northern Gannet. Weekly values were aggregated separately for waterfowl and non-waterfowl across all included species per grid cell, using mean abundance from January to March.

#### 2.1.3. Terrestrial mammal habitat data

Habitat suitability maps for eight terrestrial mammal species confirmed positive for H5N1 and with potential for contact with farmed cattle were sourced from the Gap Analysis Project (GAP) (U.S. Geological Survey (USGS), 2018). The eight species were: Abert’s Squirrel (*Sciurus aberti*), American Marten (*Martes americana*), Coyote (*Canis latrans*), Fisher (*Martes pennanti*), Red Fox (*Vulpes vulpes*), Raccoon (*Procyon lotor*), Striped Skunk (*Mephitis mephitis*), and Virginia Opossum (*Didelphis virginiana*). These were aggregated into a composite terrestrial mammal habitat density surface across the US.

#### 2.1.4. Cattle and poultry density data

County-level cattle and poultry inventory estimates were obtained from the United States Department of Agriculture National Agricultural Statistics Service (USDA-NASS) (USDA, n.d.). Cattle density was the total cattle and calves inventory per county divided by county area (km^2^); poultry density was the aggregate inventory of broiler chickens, layer chickens, pullets, roosters, ducks, geese, turkeys, and quail divided by county area. Both were log-transformed for visualisation. Counties in the top quartile of cattle density with no confirmed outbreak served as controls for the case-control analysis (n=767).

#### 2.1.5. Geographic boundary and flyway data

County-level cartographic boundary files for the contiguous US were obtained from the US Census Bureau (2023) (US Census Bureau, 2024), and migratory bird flyway boundaries from the U.S. Fish and Wildlife Service (USFWS) (United States Fish and Wildlife Service, 2015). All spatial data were projected using the World Geodetic System (WGS 1984) coordinate reference system.

### 2.2. Case-Control Ecological Exposure Analysis

To test whether the ecological conditions at outbreak counties were different from comparable non-outbreak locations, we conducted a case-control analysis. The 22 unique counties with at least one confirmed H5N1 dairy farm outbreak by 5 May 2024 were defined as cases. 42 individual outbreaks occurred across 22 counties, as several counties such as Curry County, New Mexico had multiple outbreaks. Controls were all US counties in the top quartile of national cattle density with no confirmed outbreak (n=767), restricting comparison to high-cattle-density counties so that any differences reflect conditions specific to outbreak locations rather than simply being agricultural. A comparison against all US counties, including urban and low-density rural areas, would be an unfair test of the ecological exposure.

For each county, we extracted the mean relative abundance of waterfowl (36 affected species) and non-waterfowl birds (103 affected species), and terrestrial mammal habitat density, averaged across January 2024 to the week of each county’s first confirmed outbreak, capturing exposure conditions at the time of introduction rather than a standardised seasonal average. Waterfowl abundance for the outbreak-week window was available for 18 of the 22 outbreak counties; the remaining four lacked coverage for their outbreak week and were excluded from the waterfowl comparison only. Bird abundance values were derived from eBird Status and Trends 2022 percentile-scaled rasters, normalised to a 0–1 scale, and applied as a proxy for 2024 conditions, representing expected seasonal presence of affected species rather than confirmed infection prevalence.

Differences in exposure between case and control counties were tested using the two-sided Mann-Whitney U test, chosen given the small case group (n=22), non-normally distributed bounded (0–1) exposure values, and unequal group sizes. Effect sizes were reported as the rank-biserial correlation coefficient r (negligible <0.1, small 0.1–0.3, medium 0.3–0.5, large >0.5). A Bonferroni correction for three simultaneous variables (waterfowl, non-waterfowl, terrestrial mammals) set the adjusted significance threshold at α=0.0167. County was used as the spatial unit throughout, as confirmed outbreak records were consistently available at that level.

### 2.3. Knox Space-Time Cluster Test

Space-time clustering of dairy farm outbreaks was tested using the Knox test (Knox & Bartlett, 1964), which evaluates whether event pairs close in space are also close in time more often than expected by chance. All 42 outbreak events were included, defined by confirmed start date and county centroid; events sharing a centroid were retained as separate events when start dates differed. Spatial distances were computed using the Haversine formula (Earth radius 6,371 km); temporal distances were the absolute difference in days between start dates.

The Knox statistic *K*(δ*s*, δ*t*) counted event pairs with spatial distance < δ*s* (spatial threshold) and temporal distance < δ*t* (temporal threshold), excluding self-pairs. Significance was assessed using 99,999 Monte Carlo permutations, randomly reassigning outbreak dates to spatial locations to generate a null distribution of *K* under spatial-temporal independence.

A sensitivity grid spanned six spatial thresholds (δ*s* = 25, 50, 75, 100, 150, 200 km) and five temporal thresholds (δ*t* = 3, 5, 7, 10, 14 days), yielding 30 combinations. A reference analysis used δ*s*=75 km and δ*t*=14 days, approximating intrastate cattle transport distance and estimated detection lag for H5N1 in dairy cattle. The Bonferroni-corrected threshold for 30 comparisons was α=0.0017, and the Knox ratio (observed *K* / mean null *K*) served as the effect size.

To assess whether repeat outbreaks within the same county (sharing centroid coordinates and contributing zero-distance pairs) influenced the primary analysis, a sensitivity analysis used only the first confirmed outbreak per county (n=22), providing a test unaffected by within-county repeat events.

The Knox test identifies non-random space-time clustering but does not assign a specific transmission mechanism. Significant clustering was therefore interpreted as evidence of spatial-temporal structure consistent with clustered exposure or amplification processes, rather than direct evidence of a particular pathway.

## 3. Results

A total of 139 infected wild bird species have reported cases in the US (United States Department of Agriculture Animal and Plant Health Inspection Service, 2026). Of the 139 bird species, 36 were categorised as waterfowl and 103 non-waterfowl, including domestic birds, charadriiformes and at least 17 other orders of birds (Supplementary Table S1). Many of these species migrate from southern wintering regions to northern breeding grounds.

### 3.1. Outbreak Distribution and Epidemiology

Of the 42 documented outbreaks in dairy farms as of 5 May 2024, all 42 were reported to the WAHIS system with county-specific data (Figure 1; Supplementary Table S2). The first reported outbreak was on 13 March 2024 in Castro county, Texas (Nguyen et al., 2025; WOAH, 2025). Outbreaks were confirmed across nine states and 22 unique counties. Texas accounted for the largest share with 12 outbreaks across six counties (Dallam, Castro, Hale, Hartley, Lamb, and Moore), followed by Michigan (10 outbreaks across six counties: Ionia, Montcalm, Isabella, Barry, Ottawa, and Gratiot), New Mexico (8 outbreaks, all within Curry County), Kansas (4 outbreaks across four counties: Kearny, Seward, Stevens, and Gray), Idaho (3 outbreaks, all within Cassia County), Colorado (2 outbreaks, both in Broomfield County), and single outbreaks in Ohio (Wood County), South Dakota (Brown County), and North Carolina (Wake County). The initial outbreak occurred on 10 March 2024 in Castro County, Texas, preceding the official confirmation of A/H5N1 in dairy cattle by 15 days. Of the 42 dairy farm outbreaks, 18 reported start dates prior to that official confirmation. Castro County, Texas had the third highest cattle density reported in the United States (ρ=0.14).

**Figure 1:**
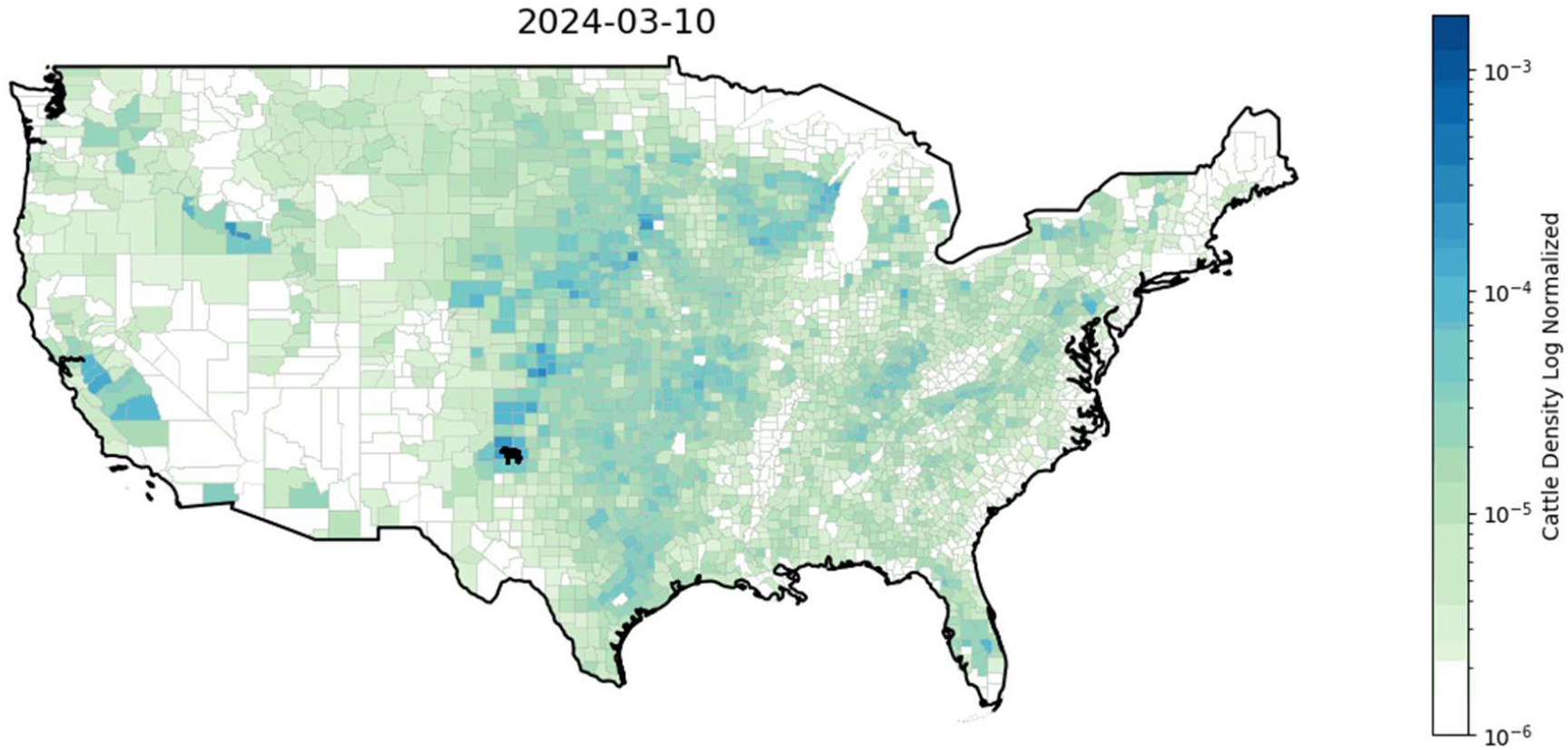
Time Series(animated) plot of the distribution of dairy farm outbreaks (n=42) layered over cattle density per county area from 10 March to 11 2024 (WOAH, 2025).

### 3.2. Exposure Analysis

We illustrate the migration pathways of wild waterfowl (Figure 2) and non-waterfowl (Figure 3) species, along with the density of habitats for the eight terrestrial mammals (Figure 4) that have tested positive for the virus since 2021. The estimated dairy cattle exposure to non-waterfowl species within the outbreak regions (ρ=0.697, z-score=1.052) exceeded that of waterfowl (ρ=0.502, z-score=0.423). In addition, the estimated dairy cattle exposure within the outbreak regions to the selected terrestrial mammals was 0.316 (z-score= -0.552).

**Figure 2:**
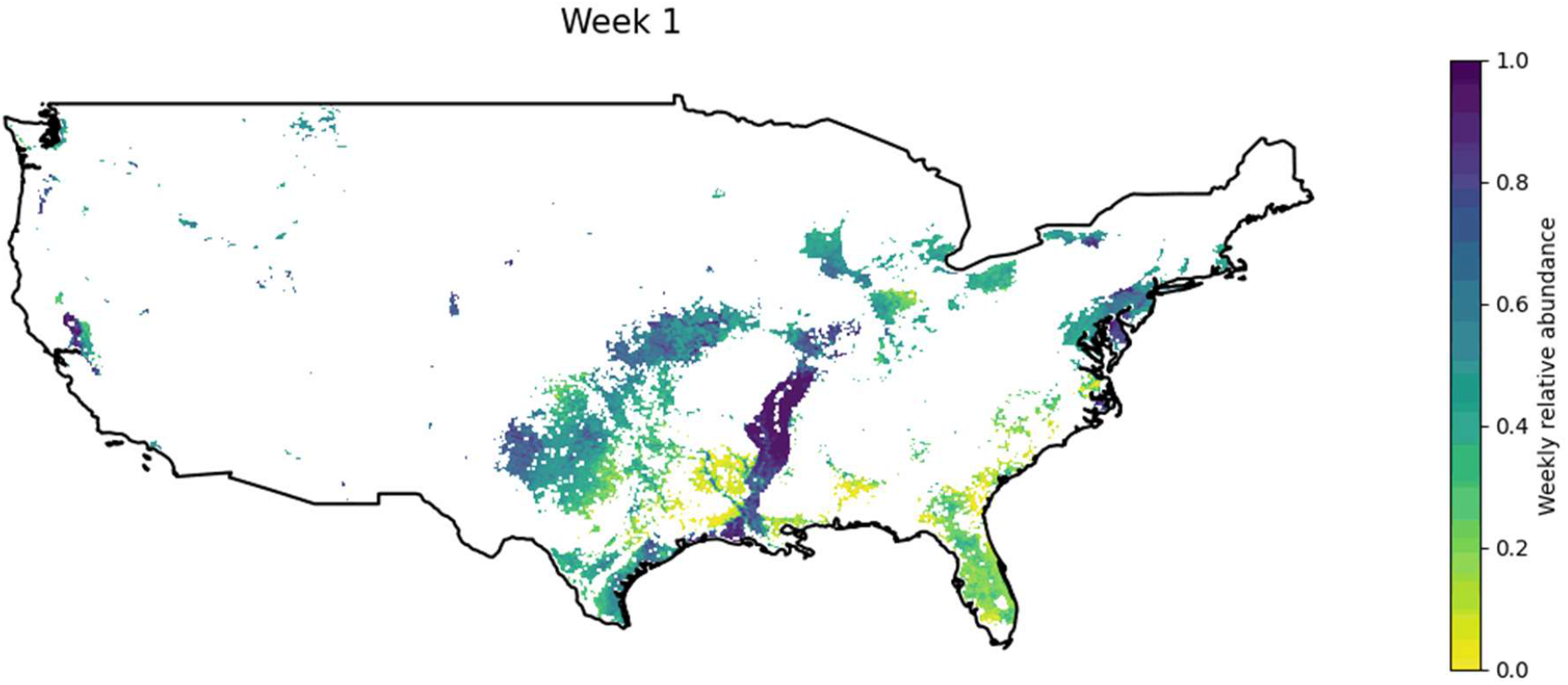
Time series(animated) map of the estimated weekly relative abundance of 36 waterfowl species affected by highly pathogenic avian influenza virus from 4 January (Week1) to 12 April (Week 14) layered with dairy farm outbreaks.

**Figure 3:**
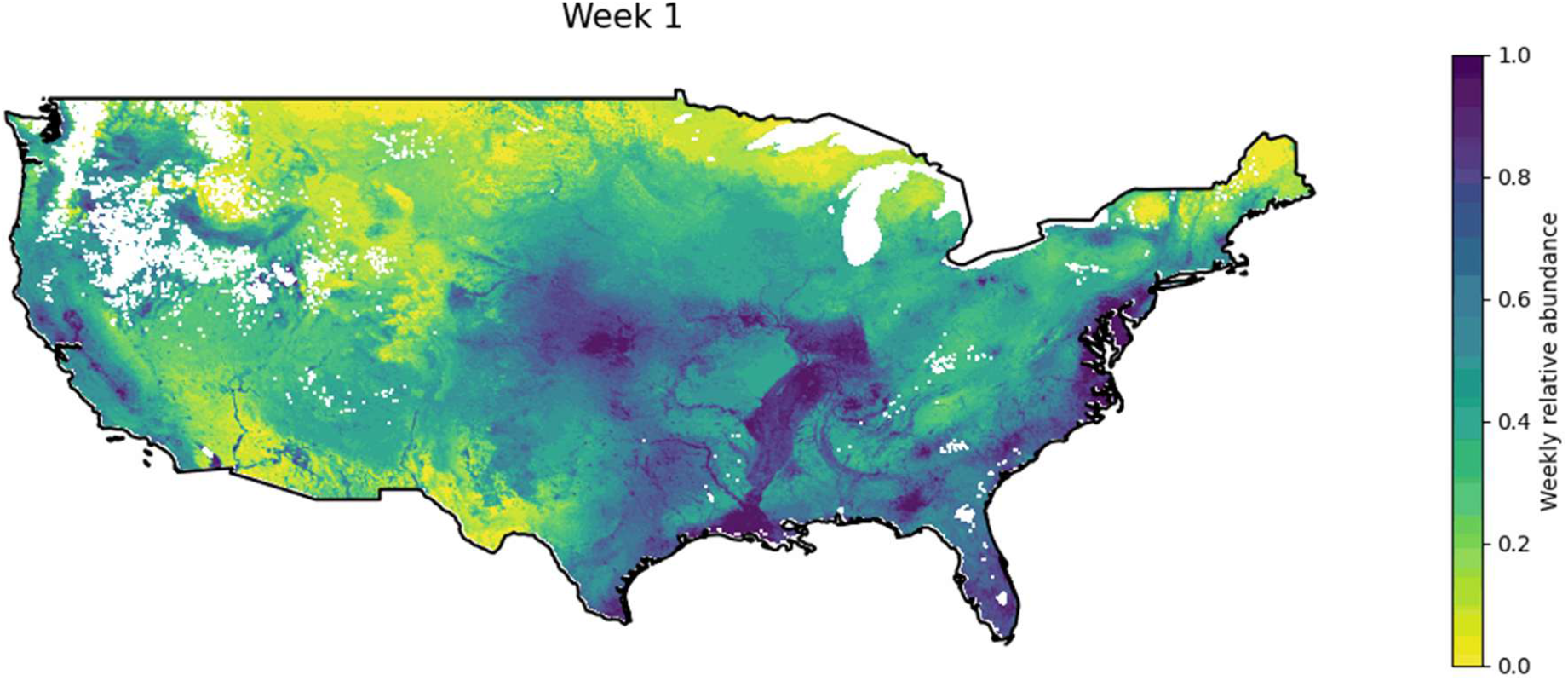
Time series(animated) map of the estimated weekly relative abundance of 103 non-waterfowl affected by highly pathogenic avian influenza virus from 4 January (Week1) to 12 April (Week 14) layered with dairy farm outbreaks.

**Figure 4:**
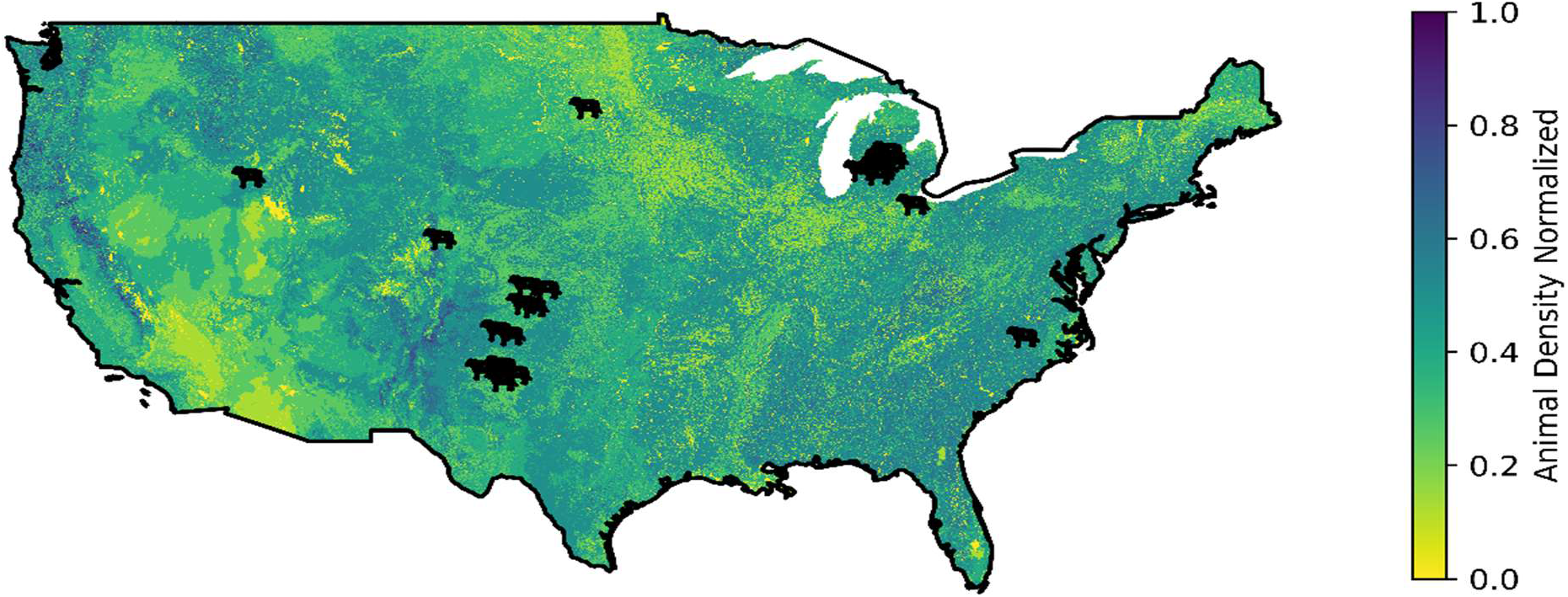
Terrestrial wild mammal density per 14km^2^ area in the mainland USA, based on potential interactions with cattle layered with dairy farm outbreaks.

In Castro County, Texas, the estimated relative abundance was 0.748 for waterfowl, 0.877 for non-waterfowl, and a terrestrial wild mammal density of 0.487 per 14km^2^. Within the subsequent three days, three more farm outbreaks emerged: one in Hale County, Texas; another in Castro County, Texas; and the third in Cassia County, Idaho. Given the geographic spread of these outbreaks over a short timeframe, we conducted an analysis of the outbreak in Cassia County, revealing estimated relative abundances of 0.654 for waterfowl, 0.749 for non-waterfowl, and a terrestrial wild mammal density of 0.375.

To formally test whether outbreak counties had significantly higher ecological exposure than comparable non-outbreak counties, we conducted a case-control analysis comparing the 22 unique outbreak counties against 767 high-cattle-density counties with no confirmed outbreak. Results are presented in Table 1. Non-waterfowl abundance was the only exposure variable significantly elevated at outbreak counties compared to controls (median 0.683 vs 0.476, p=0.00001, r=−0.541, large effect), surviving Bonferroni correction. Waterfowl abundance was not significantly different between outbreak and control counties (median 0.447 vs 0.417, p=0.271, small effect), nor was terrestrial mammal habitat density (median 0.375 vs 0.394, p=0.121, small effect).

**Table 1:** Case-control comparison of ecological exposure at H5N1 dairy farm outbreak counties (n=22) versus high-cattle-density non-outbreak control counties (n=767). Mann-Whitney U test; r = rank-biserial correlation coefficient. *p<0.00001 after Bonferroni correction (adjusted α=0.0167).

| Exposure variable | Median | Control median (n=767) | Number of cases | p-value | r |
| --- | --- | --- | --- | --- | --- |
| Waterfowl abundance | 0.447 | 0.417 | 18 | 0.271 | -0.151 |
| Non-waterfowl abundance | 0.683 | 0.476 | 22 | 0.00001* | -0.541 |
| Terrestrial mammal habitat density | 0.375 | 0.394 | 22 | 0.121 | 0.193 |

### 3.3. Space-time cluster analysis

Space-time clustering of H5N1 dairy farm outbreaks was formally tested using the Knox test with 99,999 Monte Carlo permutations across a sensitivity grid of six spatial thresholds (25-200 km) and five temporal thresholds (3-14 days), restricted to the first confirmed outbreak per county (n=22 unique counties) to avoid artificial inflation of the Knox statistic by repeat outbreaks sharing identical county centroid coordinates. Of 30 threshold combinations evaluated, 14 returned statistically significant clustering (p<0.05), and five survived Bonferroni correction for 30 comparisons (adjusted α=0.0017). P-values for the full sensitivity grid are presented in Table 2.

**Table 2:** Knox test p-values and ratios across six spatial thresholds (25-200 km) and five temporal thresholds (3-14 days). Each cell: p-value | Knox ratio. * Bonferroni-corrected significance (p<0.0017).

| Spatial (km) →<br>Temporal (days) ↓<br>Threshold | 25 km | 50 km | 75 km | 100 km | 150 km | 200 km |
| --- | --- | --- | --- | --- | --- | --- |
| 3 days | 1.000 \| 0.000 | 0.204 \| 2.273 | 0.040 \| 2.484 | 0.023 \| 2.322 | 0.013 \| 2.313 | 0.003 \| 2.356 |
| 5 days | 1.000 \| 0.000 | 0.401 \| 1.465 | 0.070 \| 1.923 | 0.030 \| 1.926 | 0.002 \| 2.320 | 0.000 \| 2.336* |
| 7 days | 0.290 \| 3.453 | 0.319 \| 1.475 | 0.066 \| 1.722 | 0.004 \| 2.011 | 0.000 \| 2.225* | 0.000 \| 2.195* |
| 10 days | 0.407 \| 2.459 | 0.577 \| 1.051 | 0.293 \| 1.228 | 0.064 \| 1.433 | 0.012 \| 1.586 | 0.000 \| 1.731* |
| 14 days | 0.524 \| 1.908 | 0.800 \| 0.817 | 0.280 \| 1.193 | 0.114 \| 1.272 | 0.039 \| 1.355 | 0.001 \| 1.518* |

No significant clustering was detected at 25 km or 50 km across any temporal window (p=0.204-1.000). At 25 km, Knox ratios of 0.00 were observed at 3-day and 5-day temporal thresholds, indicating that no outbreak pairs fell within this spatial-temporal window, while the ratio of 3.453 at 7 days did not reach significance (p=0.290). Significant space-time clustering was instead concentrated at spatial thresholds of 100-200 km, with five cells surviving Bonferroni correction, all at 150-200 km (p=0.00007-0.00114, Knox ratios 1.518-2.336). Knox ratios across the full grid ranged from 0.00 to 3.453. Clustering at these regional spatial scales was consistent with inter-county and interstate cattle transport distances which supports dispersal due to cattle trade as a potential mechanism linking geographically distinct outbreak counties.

### 3.4. Cattle-poultry outbreak interaction

A total of 30 poultry and 28 domestic outbreaks were reported in 2024, many within a 50-mile radius of dairy farm outbreaks (Figure 5). This spatial proximity was consistent across multiple states: in Texas, cattle outbreaks in Castro County (10 and 13 March 2024) preceded poultry outbreaks in neighbouring Parmer County (1 April 2024) and a backyard poultry outbreak in adjacent Deaf Smith County (13 March 2024). In New Mexico’s Curry County, four cattle outbreaks between 16 March and 2 April 2024 were followed by poultry outbreaks in neighbouring Parmer and Roosevelt counties, Texas. Similar dairy-then-poultry outbreak sequences within neighbouring counties were observed in Michigan (Montcalm, Ionia, and Isabella counties), Kansas (Kearny, Grant, and Hartley counties), and Idaho (Cassia and Twin Falls counties), with poultry outbreaks typically following cattle outbreaks by one to three weeks.

**Figure 5:**
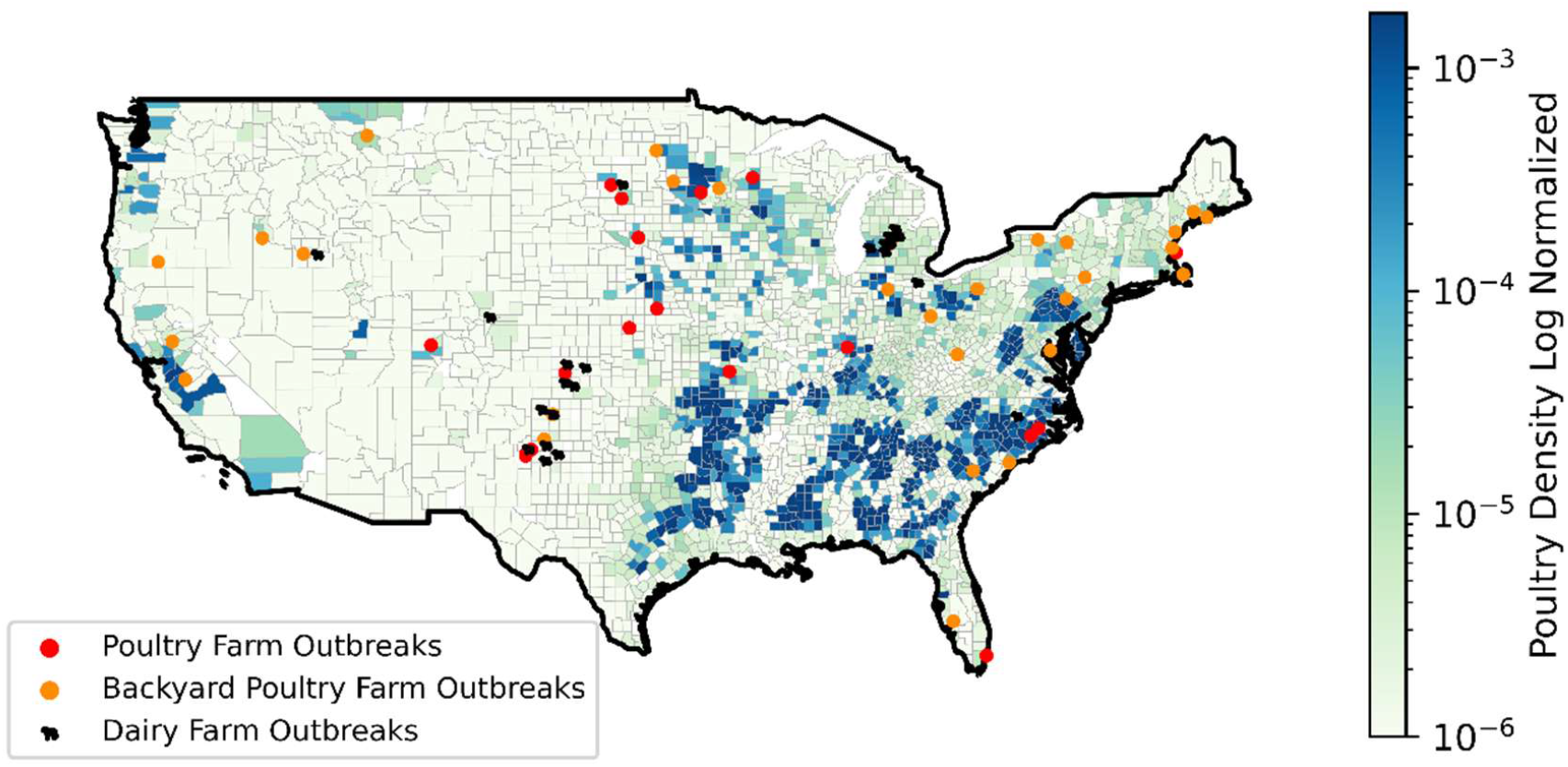
Dairy, poultry and backyard farm outbreaks in the mainland USA from January to 5 May 2024 overlayed poultry density per county area.

## 4. Discussion

Non-waterfowl abundance was as the ecological feature most strongly associated with early HPAI A/H5N1 dairy outbreak counties compared to high-cattle-density counties without reported outbreaks. Together with previous genomic evidence of a single introduction into dairy cattle, the observed space-time clustering suggests subsequent regional amplification(Caserta et al., 2024; Nguyen et al., 2025). Non-waterfowl species may therefore represent an under-recognised component of the wildlife-cattle interface, with subsequent spread shaped by regional agricultural connectivity.

The Knox test identified significant clustering at spatial thresholds of 150–200 km, whereas no significant clustering detectedat 25 km or 50 km. This regional-scale pattern is more compatible with cattle movement between counties and states than localised farm-to-farm transmission, consistent with the documented role of interstate cattle trade in amplifying the outbreak. Analysis at county centroid level may underestimate true clustering, and confirmed outbreak dates reflect reporting rather than true infection dates, which could attenuate the clustering signal. Continued outbreaks in Texas and New Mexico after spring migration shifted northward suggest that migration alone cannot explain the full outbreak sequence. Regional amplification, delayed detection, farming practicesand agricultural connectivity may also have contributed following the initial introduction into cattle. Similarly, a previous A/H5N1 and A/H5N2 in US turkey farms acould not be explained by wild-bird pathways alone (Bui et al., 2016).

The case-control analysis confirmed that non-waterfowl exposure, rather than waterfowl or terrestrial mammal habitat density, was significantly elevated at outbreak counties, indicating it is a stronger ecological correlate of early outbreak locations. Poultry outbreaks occurred near dairy farms both before and after cattle detections, suggesting shared exposure or epidemiological connectivity, although the direction of transmission could not be determined. The overlap of high cattle density, seasonal bird abundance and nearby poultry outbreaks may have created conditions favourable to initial introduction or shared exposure.Because controls were selected nationally rather than matched by flyway or state, residual differences in regional ecology and surveillance intensity may remain. Future analyses using geographically matched controls should test the strength of the non-waterfowl assosciation.

Of the 42 outbreaks, twenty occurred in Texas and New Mexico after the week of 17 March 2024, despite the highest waterfowl and non-waterfowl abundance being concentrated in the Midwest rather than the Southwest. Composite ecological-agricultural z-scores were highest in Texas counties, whereas Mighigan counties had lower scores despite high cattle density (Supplementary Figure S1). This contrast is consistent with differing contributions of ecological exposure and cattle movement across regions, but does not identify the route of introduction into individual farms.

Continual cattle trade and initially lenient trade restrictions may have contributed to interstate spread. Genomic and epidemiological investigations linked the movement of infected cattle from Texas to farms in other states, including Michigan (Sreenivasan et al., 2024; USDA APHIS, 2024). Poultry litter used as feed, bedding, or fertiliser has been proposed as a potential fomite (Sreenivasan et al., 2024; Kamel et al., 2025). However, farm-level cattle movement and poultry-litter distribution data were unavailable, so these pathways were not directly tested.

These findings support proactive surveillance at wildlife-livestock interfaces, particularly where high cattle density overlaps with substantial non-waterfowl abundance. Surveillance should integrate wild-bird, poultry and cattle data and be strengthened before seasonal migration periods to improve early detection of potential spill over risk points and regional amplification.

## 5. Conclusion

Among the ecological exposure variables examined, we identified the highest exposure to non-waterfowl avian species. Space-time cluster analysis restricted to unique outbreak counties confirmed non-random spatiotemporal structure, with significant clustering concentrated at regional spatial scales of 150-200 km. In addition, a spatial and temporal overlap between poultry and dairy outbreaks was observed descriptively, indicating potential poultry-dairy interface risk, shared exposure, or regional outbreak overlap. This highlights the threat of A/H5N1 spread through newly infected non-waterfowl wild bird species, which is a departure from the historical spread by waterfowl. These findings add spatial ecological evidence that non-waterfowl species should be considered alongside waterfowl in surveillance at wildlife-livestock interfaces. Our data support non-waterfowl abundance as a spatial correlate of early dairy outbreak counties, but further genomic, environmental, and farm-level movement data are needed to determine whether these species directly seeded outbreaks. However, additional factors such as livestock trade, poultry litter feed and contaminated milking machinery may have contributed to amplifying the epidemic. Over the past six years, the epicenter of avian influenza has shifted from Asia to Europe and the Americas, possibly due to new bird species being infected, therefore the risk of a human pandemic emerging is greatest in these new regions. This makes it imperative to understand the rapid spread in farmed ruminants in the United States.

## Data Availability

All data produced are available online:

https://doi.org/10.6084/m9.figshare.25728336

## Data, code, and materials availability

All data are available in the main text or the supplementary materials.

## Author Contribution

Conceptualization: MJ, HS, MS,

Data Collection: RD, AQ, MJ, HS

Methodology: MJ, HS, SL

Software: MJ

Investigation: MJ, HS,

Visualization: MJ, HS, SL

Funding acquisition: MJ

Supervision: SL, RM

Writing – original draft: MJ, HS,

Writing – review & editing: MJ, SL, RM

## Ethical statement

This study did not involve direct contact with, or experimentation on, live animals or human subjects. All data used were obtained from publicly available secondary sources, including confirmed disease outbreak records from the World Organisation for Animal Health’s World Animal Health Information System (WAHIS), county-level livestock inventory data from the USDA National Agricultural Statistics Service, and wild bird and terrestrial mammal distribution data from eBird Status and Trends and the U.S. Geological Survey Gap Analysis Project. As no primary animal or human data were collected, and no direct interaction with live animals or human participants occurred, institutional ethical approval was not required for this study.

## Supplementary Information

Table S1. Dairy Farm Outbreaks by associated WAHIS outbreak reference number, start date, state and county (WOAH, 2025).

Table S2. Affected wild bird species caused by highly pathogenic avian influenza H5N1 in the United States from 2021 to April 2024.

Figure S1. Composite z-score map

## References

Bui, C. M., Gardner, L., & MacIntyre, C. R. (2016). Highly Pathogenic Avian Influenza Virus, Midwestern United States. Emerging Infectious Diseases, 22(1), 138–139. 10.3201/eid2201.151053

Caserta, L. C., Frye, E. A., Butt, S. L., Laverack, M., Nooruzzaman, M., Covaleda, L. M., Thompson, A. C., Koscielny, M. P., Cronk, B., Johnson, A., Kleinhenz, K., Edwards, E. E., Gomez, G., Hitchener, G., Martins, M., Kapczynski, D. R., Suarez, D. L., Alexander Morris, E. R., Hensley, T., … Diel, D. G. (2024). Spillover of highly pathogenic avian influenza H5N1 virus to dairy cattle. Nature, 634(8034), 669–676. 10.1038/s41586-024-07849-4

Fink, D., Auer, T., Johnston, A., Strimas-Mackey, M., Ligocki, S., Robinson, O., Hochachka, W., Jaromczyk, L., Crowlye, C., Dunham, K., Andrew, Davies, I., Rodewald, A., Ruiz-Gutierrez, V., & Wood, C. (2023). eBird Status and Trends (Version 2022) [Dataset]. Cornell Lab of Ornithology, Ithaca, New York. 10.2173/ebirdst.2022

Food and Agriculture Organization of the United Nations. (2024). Bird & mammal species affected by H5Nx HPAI. FAO AnimalHealth. https://www.fao.org/animal-health/situation-updates/global-aiv-with-zoonotic-potential/bird-species-affected-by-h5nx-hpai/en

Jindal, M., Stone, H., Lim, S., & McIntyre, R. (2025). A geospatial perspective toward the role of wild bird migrations and global poultry trade in the spread of highly pathogenic avian influenza H5N1. GeoHealth, 9. 10.1029/2024GH001296

Kamel, M., Aleya, S., Almagharbeh, W. T., Aleya, L., & Abdel-Daim, M. M. (2025). The emergence of ighly pathogenic avian influenza H5N1 in dairy cattle: Implications for public health, animal health, and pandemic preparedness. European Journal of Clinical Microbiology & Infectious Diseases, 44(8), 1817–1833. 10.1007/s10096-025-05147-z

Knox, E. G., & Bartlett, M. S. (1964). The Detection of Space-Time Interactions. Journal of the Royal Statistical Society. Series C (Applied Statistics), 13(1), 25–30. 10.2307/2985220

Nguyen, T.-Q., Hutter, C. R., Markin, A., Thomas, M., Lantz, K., Killian, M. L., Janzen, G. M., Vijendran, S., Wagle, S., Inderski, B., Magstadt, D. R., Li, G., Diel, D. G., Frye, E. A., Dimitrov, K. M., Swinford, A. K., Thompson, A. C., Snekvik, K. R., Suarez, D. L., … Anderson, T. K. (2025). Emergence and interstate spread of highly pathogenic avian influenza A(H5N1) in dairy cattle in the United States. Science, 388(6745), eadq0900. 10.1126/science.adq0900

Sreenivasan, C. C., Li, F., & Wang, D. (2024). Emerging Threats of Highly Pathogenic Avian Influenza A (H5N1) in US Dairy Cattle: Understanding Cross-Species Transmission Dynamics in Mammalian Hosts. Viruses, 16(11), 1703. 10.3390/v16111703

Turnbull, M. L., Zakaria, M. K., Upfold, N. S., Bakshi, S., Magill, C., Das, U. R., Clarke, A. T., Mojsiejczuk, L., Herder, V., Dee, K., Liu, N., Folwarczna, M., Ilia, G., Furnon, W., Schultz, V., Chen, H., Devlin, R., McCowan, J., Young, A. L., … Palmarini, M. (2025). The potential of H5N1 viruses to adapt to bovine cells varies throughout evolution. Nature Communications, 16, 11042. 10.1038/s41467-025-67234-1

United States Department of Agriculture Animal and Plant Health Inspection Service. (2026, May 22). Detections of Highly Pathogenic Avian Influenza in Wild Birds. USDA APHIS. https://www.aphis.usda.gov/livestock-poultry-disease/avian/avian-influenza/hpai-detections/wild-birds

United States Fish and Wildlife Service. (2015). USFWS Administrative Waterfowl Flyway Boundaries [Dataset]. https://data.cnra.ca.gov/dataset/usfws-administrative-waterfowl-flyway-boundaries

US Census Bureau. (2024). Cartographic Boundary Files—Shapefile [Dataset]. https://www.census.gov/geographies/mapping-files/time-series/geo/carto-boundary-file.html

U.S. Geological Survey (USGS), G. A. P. (GAP). (2018). U.S. Geological Survey—Gap Analysis Project Species Habitat Maps CONUS_2001 [Dataset]. U.S. Geological Survey. 10.5066/F7V122T2

USDA. (2025a). Epidemiological Bulletin: 2024 Highly Pathogenic Avian Influenza (H5N1) Livestock Event – Nevada and Arizona H5N1 D1.1 HPAI Dairy Investigation. https://www.aphis.usda.gov/sites/default/files/20250611-epi-brief-nevada-arizona-h5n1-dairy.pdf

USDA. (2025b). The Occurrence of Another Highly Pathogenic Avian Influenza (HPAI) Spillover from Wild Birds into Dairy Cattle. https://www.aphis.usda.gov/sites/default/files/dairy-cattle-hpai-tech-brief.pdf

USDA. (2026, March 13). HPAI Confirmed Cases in Livestock | APHIS. https://www.aphis.usda.gov/livestock-poultry-disease/avian/avian-influenza/hpai-detections/hpai-confirmed-cases-livestock

USDA. (n.d.). Quick Stats [Ad-hoc query tool] [Computer software]. National Agricultural Statistics Service. https://quickstats.nass.usda.gov/

USDAAPHIS. (2024, June 9). 2024 Highly Pathogenic Avian Influenza (H5N1)—Michigan Dairy Herd and Poultry Flock Summary. USDA APHIS. https://www.aphis.usda.gov/sites/default/files/hpai-h5n1-dairy-cattle-mi-epi-invest.pdf

WOAH. (2025). World Animal Health Information System (WAHIS) [Dataset]. World Organisation for Animal Health (WOAH). https://wahis.woah.org/#/event-management

