## Supplementary Materials for "Geospatial evidence of introduction of highly pathogenic avian influenza into dairy farms in the United States"

**Table S1:** Dairy Farm Outbreaks by associated WAHIS outbreak reference number, start date, state and county (WOAH, 2025).

| Outbreak Reference | Start Date | State | County |
| --- | --- | --- | --- |
| TX 011 | 2024-03-10 | Texas | Castro |
| TX 007 | 2024-03-11 | Texas | Hale |
| ID 001 | 2024-03-13 | Idaho | Cassia |
| TX 001 | 2024-03-13 | Texas | Castro |
| TX 004 | 2024-03-14 | Texas | Moore |
| TX 003 | 2024-03-14 | Texas | Hale |
| TX 010 | 2024-03-15 | Texas | Dallam |
| NM 002 | 2024-03-16 | New Mexico | Curry |
| TX 008 | 2024-03-17 | Texas | Dallam |
| KS 002 | 2024-03-20 | Kansas | Kearny |
| MI 001 | 2024-03-20 | Michigan | Montcalm |
| TX 002 | 2024-03-20 | Texas | Hartley |
| KS 001 | 2024-03-21 | Kansas | Seward |
| TX 005 | 2024-03-21 | Texas | Lamb |
| KS 004 | 2024-03-22 | Kansas | Stevens |
| KS 003 | 2024-03-25 | Kansas | Gray |
| TX 009 | 2024-03-25 | Texas | Dallam |
| OH 001 | 2024-03-27 | Ohio | Wood |
| NM 003 | 2024-03-28 | New Mexico | Curry |
| NM 004 | 2024-03-29 | New Mexico | Curry |
| SD 001 | 2024-03-29 | South Dakota | Brown |
| MI 002 | 2024-03-30 | Michigan | Montcalm |
| NM 006 | 2024-04-01 | New Mexico | Curry |
| NM 005 | 2024-04-01 | New Mexico | Curry |
| NM 007 | 2024-04-02 | New Mexico | Curry |
| TX 012 | 2024-04-03 | Texas | Hartley |
| NC 001 | 2024-04-04 | North Carolina | Wake |
| MI 004 | 2024-04-08 | Michigan | Ottawa |
| MI 005 | 2024-04-10 | Michigan | Isabella |
| MI 003 | 2024-04-11 | Michigan | Ionia |

**Table S2:** Affected wild bird species caused by highly pathogenic avian influenza H5N1 in the United States from 2021 to April 2024.

| Category | Species |
| --- | --- |
| Waterfowl | American Black Duck, American Green-winged Teal, American Wigeon, Blue-winged Teal, Bufflehead, Cackling Goose, Canada Goose, Canvasback, Cinnamon Teal, Common Eider, Common Goldeneye, Common Loon, Common Merganser, Fulvous Whistling Duck, Gadwall, Greater Scaup, Greater White-fronted Goose, Hooded Merganser, Lesser Scaup, Mallard, Mallard/Black Duck Hybrid, Mottled Duck, Muscovy Duck, Mute Swan, Northern Pintail, Northern Shoveler, Redhead Duck, Ring-necked Duck, Ross's Goose, Ruddy Duck, Sandhill Crane, Snow Goose, Trumpeter Swan, Tundra Swan, White-winged Scoter, Wood Duck. |
| Non-waterfowl | American Coot, American Crow, American Kestrel, American Robin, American White Pelican, American Wood Stork, Arctic Tern, Bald Eagle, Barn Owl, Barred Owl, Black Skimmer, Black Turnstone, Black Vulture, Black-bellied Plover, Black-billed Magpie, Black-crowned Night Heron, Black-legged Kittiwake, Boat-tailed Grackle, Bonaparte's Gull, Brandt's Cormorant, Brant, Broad-winged Hawk, Brown Pelican, California Condor, California Gull, California Quail, Caspian Tern, Cattle Egret, Common Grackle, Common Murre, Common Raven, Common Tern, Cooper's Hawk, Crested Caracara, Dark-eyed Junco, Double-crested Cormorant, Dunlin, Eared Grebe, Eastern Screech Owl, Ferruginous Hawk, Fish Crow, Forster's Tern, Gannet, Glaucous Gull, Glaucous-winged Gull, Glossy Ibis, Golden Eagle, Great Black-backed Gull, Great Blue Heron, Great Egret, Great Horned Owl, Greater Sage Grouse, Great-tailed Grackle, Green Heron, Harris Hawk, Herring Gull, Horned Grebe, House Sparrow, Laughing Gull, Long-eared Owl, Merlin, Neotropic Cormorant, Northern Fulmar, Northern Gannet, Northern Harrier, Osprey, Pacific Loon, Parasitic Jaeger, Peregrine Falcon, Pied-billed Grebe, Prairie Falcon, Red-necked Grebe, Red-necked Phalarope, Red-shouldered Hawk, Red-tailed Hawk, Red-winged Blackbird, Ring-billed Gull, Roseate Spoonbill, Rough-legged Hawk, Royal Tern, Ruddy Turnstone, Ruffed Grouse, Sabine's Gull, Sanderling, Sandwich Tern, Sharp-shinned Hawk, Short-billed Gull, Short-eared Owl, Short-tailed Shearwater, Snowy Egret, Snowy Owl, Snowy Plover, Swainson's Hawk, Thayer's Gull, Tree Swallow, Turkey Vulture, Western Grebe, Western Gull, Western Sandpiper, Western Screech Owl, White Ibis, Wild Turkey, Willet. |

**Composite z-score map**

Composite z-score represents the mean of monthly z-scores for non-waterfowl abundance (January to outbreak week) and cattle density, computed relative to national background distributions. Darker red indicates counties with higher combined ecological exposure and agricultural density. Counties are coloured only where outbreak data were available.

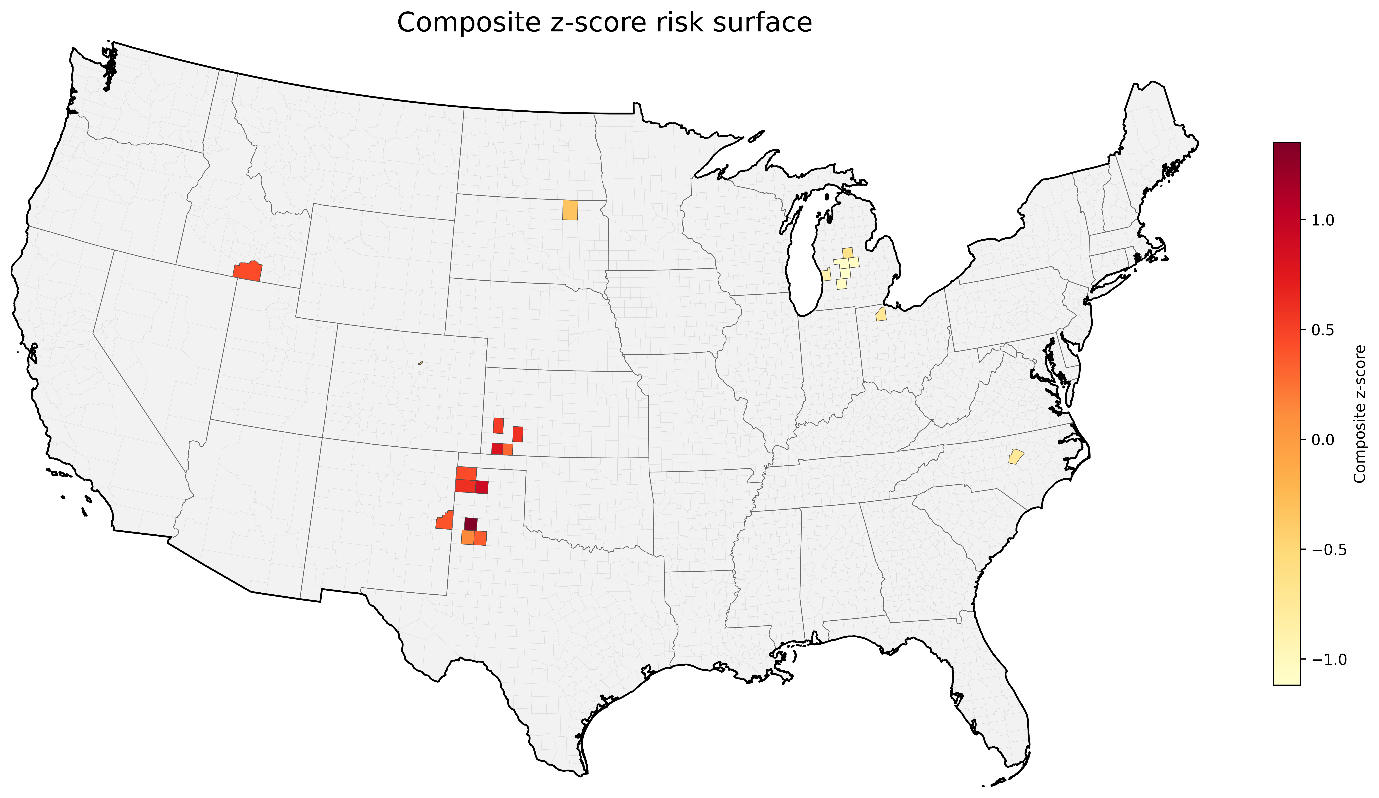

**Figure S1:** Composite ecological-agricultural z-score at confirmed H5N1 dairy farm outbreak counties (n=22 unique counties), United States, 2024.
